# Community pharmacists’ knowledge and preparedness to participate in the fight against Coronavirus disease 2019 (COVID-19) in Zambia

**DOI:** 10.1101/2020.09.01.20185694

**Authors:** Jimmy Mapenzi Hangoma, Steward Mudenda, Mpande Mukumbwa Mwenechanya, Aubrey Chichoni Kalungia

## Abstract

**Background:** Coronavirus disease (COVID-19) pandemic has severely strained healthcare systems globally. Community pharmacists remain vital health professionals with unique roles in responding to symptoms, supplying medicines, and providing health promotion in the communities where they serve. The purpose of this study was to assess the level of knowledge and preparedness of community pharmacists in Zambia as frontline workers in curbing COVID-19.

**Method:** This descriptive cross-sectional survey was conducted among 300 registered community pharmacists in Zambia. A self-administered online questionnaire was used to collect the data. Descriptive statistics were used to analyse the data using the Statistical Package for Social Sciences version 23.

**Results:** From a total of 393 community pharmacists, 300 participated in the study indicating a response rate of 76.3%. 97% of the pharmacists knew the common signs and symptoms of COVID-19, 83% knew the national guidelines for screening criteria, and 93% were aware that a suspected case of COVID-19 presenting to the community pharmacy needed to be alerted to the national response team. Only 59% knew they needed to self-isolate and self-report should they experience symptoms of COVID-19. 85% stated that personal protective equipment was inadequate in the pharmacies they operated from. 60% of the pharmacists were prepared to participate in the frontline fight against COVID-19.

**Conclusion:** Community pharmacists in Zambia are knowledgeable about COVID-19 signs and symptoms, including the technical guidelines on the screening and detection of suspected cases. Community pharmacists are prepared to participate in the frontline fight against the COVID-19 pandemic in Zambia.

## Background

There is a fatal outbreak of coronavirus disease 2019 (COVID-19) caused by a novel coronavirus named severe acute respiratory syndrome coronavirus 2 (SARS-CoV-2) [1]. The first case of COVID-19 was recorded in Wuhan, Hubei Province, China, in December 2019 and was said to be pneumonia of unknown cause [2]. The disease has since spread across many countries and the World Health Organization (WHO) declared the disease a global pandemic in February 2020 [3]. Over 5,590,358 cases of COVID-19 infections have been confirmed globally and over 347,907 mortalities have since been recorded globally [4]. During the time this study was being conducted, the most affected countries were China, Italy, France, Spain, the United Kingdom, and the USA [5]. In African countries, however, the number of cases has been on the rise sharply and the negative socio-economic and health effects of the pandemic are estimated to be severe due to relatively poor health care systems and small economies in most of the countries. As the pandemic began to take its toll on Africa, it’s devastating impact was anticipated [6, 7].

In Zambia, the first confirmed cases of COVID-19 were reported on 18^th^ March 2020, and since then, the number of confirmed cases has been increasing steadily [8]. In order to curb the spread of the disease, a scalable national response has seen the activation of a public health emergency response mechanism spearheaded by the Ministry of Health through the Zambia National Public Health Institute (ZNPHI) [9]. Key activities in this response include enhancing disease surveillance, health human resource capital development, case detection and management in designated public healthcare facilities, restrictions on social gatherings and non-essential travel, disease prevention, and health promotion among other strategies [9].

The rate at which the disease was spreading globally, WHO feared that most health systems would be strained particularly in Africa, and thus there was a need for a multisectoral action to find containment solutions. One of the key sectors to consider as part of the solutions was the community pharmacies [10]. According to the International Pharmaceutical Federation (FIP), pharmacists and the pharmacy workforce at community and hospital pharmacies and clinical biology laboratories are capable of preventing the spread of the new coronavirus disease, advising the public, and supporting the efficient management of infection by healthcare systems. Since pharmacies are often the first point of contact with the health system, the FIP made a call to national governments to support the sector in these roles and to count on pharmacists worldwide as key partners in the global effort to contain the outbreak [10].

Community pharmacists were usually the first points of contact for members of the community seeking primary health care services and access to medicines and pharmaceutical products in Zambia [11, 12]. With cases of COVID-19 on the rise in Zambia, there was an urgent need for health system managers to enhance the national response by maximising the use of currently available health workforce and resources. It is therefore vital at this time of the pandemic that existing services are comprehensively reviewed and fully utilise any unrealised potential among healthcare providers across the continuum of care. Community pharmacy is one of a number of health professions that has a key role to play in responding to the current pandemic. As the scope of community pharmacy practice varies considerably across countries, it is important to examine ways in which the profession can assist with the public health response to COVID-19 and maintaining the continuity of healthcare services [13].

Other countries have enhanced this sector’s participation in the fight against the pandemic to varying degrees. In Taiwan, the government, in their national response had to allow all hospitals, clinics, and pharmacies to have access to a patients’ travel history in order to curb the spread of the deadly infection [14]. Community pharmacists can reinforce mitigation behaviours by applying the health belief model (HBM) [15]. This commentary provides an overview of the HBM and offers suggestions on how community pharmacists can use it as a guide to patient communication in these uncertain contexts [15]. Putt has stressed that people may not think of a pharmacist as an essential healthcare worker because they do not necessarily see them running around the emergency room at their local hospital, but these health care providers are more essential than people might think [16]. Pharmacists are working on the frontline to ensure patient safety while putting their lives at risk every single day. In the current situation with the COVID19 pandemic, community pharmacists work as frontline workers but receive less support including inadequate personal protective equipment (PPE) [10]. Anyone can walk up to a pharmacy counter, making the pharmacist the most accessible health care worker to patients across the country, but also putting the pharmacist at the highest risk for contracting the virus [16]. As stated by the FIP, community pharmacists have a key role in preventing the spread of SARS-CoV-2 [10]. They are charged with key responsibilities of informing, advising and educating the community, maintaining a stable supply of pharmaceuticals and personal hygiene products, and screening of suspected cases of COVID-19, and making the appropriate referral as required [10]. Consequently, community pharmacists can contribute to the early detection and appropriate referral of possible cases of the COVID-19, with the goal to contribute to the prevention of the overall community spread of the virus in Colombia [17]. Rural communities are often isolated from large health centres with the capabilities of handling an outbreak such as COVID-19. Rural patients also face unique challenges such as extended travel time to an acute care facility, hazardous terrain, and the lack of reliable or public transportation. These geographic challenges may cause patients to postpone or go without accessing needed health services. Amidst the avalanche of reports concerning the spread of the virus, there is also recognition that pharmacists are in a unique position to make essential public health contributions, especially in rural areas, to address the shortage of primary care providers, nurses, and physicians [18].

Community pharmacists, therefore, need to position themselves to not only provide pharmaceutical services but further prevent the spread of the virus thereby contributing greatly to combating the pandemic in Zambia. As a starting point, we assessed the preparedness of pharmacists serving in the community retail pharmacy sector to participate in the frontline national response to COVID-19 in Zambia. Specifically, we determined their knowledge about COVID-19, technical guidelines for case detection, and screening, including availability and use of PPE in the pharmacy. This information could help to inform future decisions about the restructuring of existing health services by governments, public health bodies, and policymakers in response to public health crises such as COVID-19. This information could help inform health system managers’ future decisions and considerations about prudent use of the available health workforce, restructuring of existing health services, and response strategies by ZNPHI and other state actors involved in the response to public health emergencies such as COVID-19.

## Methods

### Study design

This was a descriptive cross-sectional survey involving **c**ommunity pharmacists providing pharmaceutical services in retail pharmacy outlets across Zambia. This study was conducted among community pharmacists because they are among the first point of contact with patients in the communities.

### Sample size and sampling

The sample size was purposive and included all community pharmacists in registered pharmacies as at close of 2019. The total number of registered pharmacies in the country was obtained from Zambia Medicines Regulatory Authority (ZAMRA) at the time of this study was 393 [19]. As per statutory requirements, each registered community retail pharmacy is managed by a registered pharmacist. The response rate was 300 out of 393 giving a percentage response rate of 76.3%.

### Data collection tool and procedures

An online questionnaire with closed-ended questions was sent to selected community pharmacists using Google forms. The questionnaire was pre-tested by administering it to 10 community pharmacists in Lusaka and it was validated for consistency. The questionnaire was used to collect quantitative data on participants’ knowledge of COVID-19 symptoms, prevention strategies, national recommended screening and testing protocols, level of preparedness to participate in the frontline, and use of personal protective equipment.

### Data analysis

Descriptive statistics were used to analyse the data using the Statistical Package for Social Sciences version 23. Frequencies and proportions were used to describe the data. The findings were presented in form of tables.

### Ethical Considerations

Ethical approval was granted by the University of Zambia Health Sciences Research Ethics Committee (UNZAHSREC) with an Ethics Approval of Protocol ID Number 20203101007.

## Results

Table 1 shows the demographic information of the participants. The mean (SD) age of participants was 32.30 (+/-5.88) years, the majority of whom were males (n=210) and the majority being married (n=170). All the participants had attained bachelors’ degrees (pharmacists), mean work experience (SD) of 7.6 (+/-5.06) years.

**Table 1:** Demographic Information of the Participants.

| **Characteristic** | **Frequency**<br>**(Number)** | **Percent (%)** |
| --- | --- | --- |
| **Age in years, mean (SD)** | 32.30 ($\pm 5.88$) | |
| **Sex** |  |  |
| Male | 210 | 70 |
| Female | 90 | 30 |
| **Marital Status** |  |  |
| Single | 130 | 43 |
| Married | 170 | 57 |
| Divorced | 0 | 0 |
| Widowed | 0 | 0 |
| **Level of Education** |  |  |
| Bachelor's | 231 | 77 |
| Master's | 51 | 17 |
| Other | 21 | 7 |
| **Work experience, mean (SD)** | 7.6 (5.06) |  |

Among the 300 community pharmacists that responded, the majority (97%, n=291) adequately knew the common signs and symptoms of COVID-19, 83% (n=249) knew the national guidelines for COVID-19 screening criteria, and 93% (n=279) were aware that a suspected case of COVID-19 presenting to the pharmacy needed to be alerted to the Ministry of Health or the national response team. Only 59% (n=177) knew they also needed to self-isolate and self-report should they experience symptoms of COVID-19 (Table 2). About 76% (n=228) indicated the availability of personal protective equipment (PPE) in the pharmacy while 85% (n=255) stated that PPE was insufficient in their pharmacies (Table 3). Generally, preventive measures were being implemented in community pharmacies (Table 4). The majority (87%, n=261) were able to provide initial screening and referral of suspected cases of COVID-19. Overall, community pharmacists were prepared to participate in the frontline fight against the COVID-19 pandemic in Zambia (Table 5). However, PPE was inadequate in the majority of the community pharmacies.

**Table 2:** Knowledge of Participants about COVID-19.

| **Characteristic** | **Frequency**<br>**(n=300)** | **Percent (%)** |
| --- | --- | --- |
| **Symptoms & Signs** |  |  |
| Cough | 291 | 97 |
| Fever | 291 | 97 |
| Tiredness | 190 | 63 |
| Difficulties in breathing | 291 | 97 |
| **Risks for severe symptoms & signs** |  |  |
| Older patients | 300 | 100 |
| Hypertensive patients | 231 | 77 |
| Cardiac patients | 261 | 87 |
| Diabetic patients | 270 | 90 |
| Asthmatic patients | 300 | 100 |
| **Awareness of national guidelines for COVID-19** |  |  |
| Aware | 250 | 83 |
| Not aware | 50 | 17 |
| **Screening criteria for COVID-19** |  |  |
| Anyone with a fever/respiratory infection | 288 | 96 |
| Travel in the past 14 days from a country of local transmission | 300 | 100 |
| Close contact in the past 14 days with confirmed COVID-19 infection | 300 | 100 |
| Severe respiratory infection | 267 | 89 |
| **Pharmacists' action of a suspected client of COVID-19** |  |  |
| Azithromycin+/-Hydroxychloroquine | 0 | 0 |
| Treat presenting complaints/refer the patient to the hospital | 123 | 41 |
| Tell the client to go to the nearest hospital | 135 | 45 |
| Get full details, isolate & inform MOH/response team | 216 | 72 |
| **The action of pharmacy staff in case COVID-19 case** |  |  |
| Self-isolate | 177 | 59 |
| Contact the MoH/response team | 279 | 93 |
| Knock off & come following day | 0 | 0 |

**Table 3:** Availability and Types of Personal Protective Equipment for Staff.

| **Characteristic** | **Frequency**<br>**(n=300)** | **Percent**<br>**(%)** |
| --- | --- | --- |
| **Availability of PPE** |  |  |
| Available | 228 | 76 |
| Not available | 72 | 24 |
| **Sufficiency of PPE availability** |  |  |
| Sufficient | 45 | 15 |
| Insufficient | 255 | 85 |
| **Types of PPE Available to Staff** |  |  |
| Gloves | 240 | 80 |
| Foot protective wear | 9 | 3 |
| Eye protective goggles | 69 | 23 |
| Full body gowns | 0 | 0 |
| Aprons | 39 | 13 |
| Face masks | 300 | 100 |

**Table 4:** Preventive Measures towards COVID-19 Pandemic.

| **Characteristic** | **Frequency**<br>**(n=300)** | **Percent (%)** |
| --- | --- | --- |
| Hand washing practice | 290 | 97 |
| Use of alcohol hand rubs/hand sanitizers | 279 | 93 |
| Use of face masks | 300 | 100 |
| The staff that used gloves | 114 | 38 |
| The staff that used protective gowns | 9 | 3 |
| The practice of social distancing | 279 | 93 |
| Pharmacies with phone use restrictions | 60 | 20 |
| Swabbing of swiping machines | 117 | 39 |
| Labelled floors for social distance | 210 | 70 |
| Availability of hand washing services | 300 | 100 |
| **Types of face masks used by staff** |  |  |
| N95 | 9 | 3 |
| FFP1 | 39 | 13 |
| FFP3 | 21 | 7 |
| Surgical face masks | 279 | 93 |
| Cloth face masks | 60 | 20 |
| Others | 21 | 7 |

**Table 5:** Preparedness of Community Pharmacists to participate in the fight against COVID-19 Pandemic.

| **Characteristic** | **Frequency**<br>**(n=300)** | **Percent (%)** |
| --- | --- | --- |
| Preparedness of community pharmacists to participate in the national response to COVID-19 pandemic | 180 | 60 |
| The ability of community pharmacists to provide initial screening and referral of a suspected case of COVID-19 | 261 | 87 |

## Discussion

Coronavirus disease 2019 (COVID-19) is a public health disaster that started in China and spread to the rest of the world [20]. In curbing this public health problem, the FIP has made several recommendations that community pharmacists can utilize [10]. This study focussed on the knowledge and preparedness of community pharmacists in the fight against COVID-19 as frontline workers.

### Statement of principal findings

Overall, community pharmacists have appropriate knowledge and well prepared to participate in the frontline fight against COVID-19 pandemic in Zambia. Unfortunately, PPE was inadequate in most of the community pharmacies. This challenge will require to be addressed to avoid exposing the community pharmacists to COVID-19. The scope of community pharmacy practice in Zambia enables pharmacists to be utilised in the health response to the COVID-19 pandemic. It is therefore prudent for health authorities to consider ways in which community pharmacists can be effectively utilised in the public health response to COVID-19. This will ensure that access to medicines by communities is maintained in the continuum of care during this difficult time.

Pharmacists in Zambia being highly knowledgeable and capable could be key partners in the multisectoral fight against the spread of COVID-19 infection as recommended by the FIP and WHO [10, 17, 21]. With the health system in Zambia already strained just like in many African countries, community pharmacists could offer screening, testing, and referral services for suspected or confirmed cases of COVID-19 infection. This is also in tandem with other research that reported the role of pharmacists in public health emergencies such as COVID-19 [22]. Also, during this pandemic, pharmacists should be allowed to test and treat certain conditions and perform certain acts independently of a physician such as supplying medication for chronic illnesses like HIV, asthma, hypertension, and diabetes without requesting clients for a prescription [23]. Such legislation would reduce the burden on healthcare facilities and primary care providers, nurses, and physicians as well as patients, especially those living in remote and rural areas [18].

Community pharmacists, being the first contact point for patients in communities, were at a greater risk of contracting the virus due to inadequate PPE and infection control. Though generally available, PPE was limited mainly to surgical masks and gloves. Pharmacies lacked protective gowns, foot protective wear, and eye-protective goggles. Pharmacists reported frequent hand washing especially after attending to clients, using alcohol-based hand rubs or hand sanitizers, and wearing surgical face masks. They did not, however, wear gloves, restrict the use of phones and swab swiping machines which could be routes of infection transmission. Social distance was practised despite a good number of them not having labelled the floor. Those that had labelled the floor used the recommended minimum distance of 1 metre. Similarly, Putt stated that in the current situation with the COVID-19 pandemic, pharmacists and pharmacy technicians were pumping out much more volume with much less support, including proper PPE. Despite the challenges stated above, community pharmacists must learn how to protect themselves from being exposed to and infected with COVID-19 [24]. Anyone could walk up to a pharmacy counter, making the pharmacist the most accessible health care worker to patients globally, but also putting the pharmacist at the highest risk for contracting the virus [16]. Sheppard and Thomas stated that, with the emergence of the novel coronavirus pandemic, there was a need for healthcare providers to reinforce behaviours that limited the spread of the pandemic which included social distancing and remaining in the homes whenever possible [15]. They further stated that community pharmacists could reinforce mitigation behaviours by applying the health belief model (HBM) [15]. FIP also charges that community pharmacists had a key role in preventing the spread of the 2019-CoV virus [10]. They were charged with key responsibilities of informing, advising, and educating the community and maintaining a stable supply of pharmaceuticals and personal hygiene products [17].

Given the preparedness and willingness of community pharmacists, governments have the opportunity to include them in risk communication and community engagement. Community pharmacists are frontline workers in the fight against the COVID-19 pandemic because they interact face-to-face with patients in the provision of pharmaceutical services [25]. In most cases, community pharmacists may not be seen as healthcare professionals despite their importance in the provision of healthcare services [26]. Putt stressed out that pharmacists were more essential than most people thought [16]. He pointed out that pharmacists were working on the front lines to ensure patient while putting themselves at risk every single day [16]. In Taiwan, as a result of the pressure that was mounting up due to COVID-19, the government had to allow all hospitals, clinics, and pharmacies to have access to patients’ travel history in order to curb the spread of the deadly infection [14]. Cadogan and Hughes have also stressed out the need to incorporate community pharmacists as they had a key role to play in responding to the current pandemic. They noted that the scope of community pharmacy practice varied considerably across countries, hence there was a need to examine ways in which the profession could assist with the public health response to COVID-19 and maintaining the continuity of healthcare services [13]. Information coming from community pharmacists can reinforce behaviours that limited the spread of the pandemic which included social distancing and remaining at home whenever possible. However, community pharmacists may need a quick course on health behaviour change theories, for example, the health belief model (HBM) [15], to identify what motivates people to adopt mitigation behaviours in their communities. Such an informed approach can help community pharmacists effectively discharge their key responsibilities of informing, advising, and educating the community while maintaining a stable supply of pharmaceuticals and personal hygiene products [17, 27]. Community education plays a vital role in the prevention and curbing of diseases such as COVID-19 [28, 29]. Community pharmacists are ready to deal with COVID19 based on their awareness and roles [30]. Therefore, community pharmacy practice should be strengthened and utilized in the fight against COVID-19.

### Strengths of the study

This study is the first study conducted in Zambia on the knowledge and preparedness of community pharmacists as frontline workers in the fight against COVID-19. In addition, from literature, it is among the few studies realizing the importance of community pharmacists as cardinal practitioners in the fight against COVID-19. The study was conducted country-wide in Zambia, this means that the results can be generalized to represent all community pharmacists registered and practicing in Zambia.

### Implications on policy and practice

If these findings are taken into consideration, they would help address the workforce gaps by enabling community pharmacists to actively participate in the frontline national response to COVID-19. This also presents as an opportunity to scale up screening and detection of possible COVID-19 cases and there would be continued pharmaceutical care services through the supply of medicines to clients who may not have an opportunity to access medical care.

### Limitations of the study

This was an online-based cross-sectional survey that utilised a structured questionnaire. The structured questionnaire may lead to a form of bias and hence affect the study findings. This study cannot be generalized among hospital pharmacists as it only focussed on community pharmacists.

### Unanswered questions and further research

We ask ourselves certain questions: What are the psychological impacts of COVID-19 among community pharmacists? Will the Ministry of Health engage community pharmacists as frontline workers in the fight against COVID-19? Therefore, these questions remain unanswered and will thus form the basis of future research with regards to pharmacists and COVID-19.

## Conclusion

Community pharmacists in Zambia are knowledgeable about COVID-19 signs and symptoms, including the technical guidelines on the screening and detection of suspected cases. They are prepared to participate in the frontline fight against the COVID-19 pandemic in Zambia. However, personal protective equipment was inadequate in the community pharmacies. Due to the inadequate PPE, the community pharmacists should learn how to prevent exposure to and infection with COVID-19.

What is known about this topic?
- COVID-19 affects everyone and requires a multisectoral approach in order to curb it
- Community pharmacists are the first point of contact with members of the community that require access pharmaceutical services

What this research adds
- Community pharmacists must be empowered to work as frontline workers in the fight against COVID-19
- Community pharmacists must be prepared to fight COVID-19

## Data Availability

Data is available in case other scholars want to use it

## Acknowledgements

We would like to thank all the community pharmacists who participated in this survey.

## Authors’ contributions

All authors conceptualized and designed the study. JMH, MMM, and ACK conducted data collection. JMH and SM conducted data analysis. All authors made sure that the results were interpreted. JMH drafted the initial draft of the manuscript. All authors critically revised the manuscript for important intellectual content. All authors read and approved the final version of the manuscript.

## References

1. Wilder-Smith A, Chiew CJ, Lee VJ. Can we contain the COVID-19 outbreak with the same measures as for SARS? Lancet Infect Dis. 2020; S1473-3099(20)30129-8

2. Singhal T. A review of coronavirus disease-2019 (COVID-19). The Indian Journal of Pediatrics. 2020; 87: 281–286

3. Sohrabi C, Alsafi Z, O’Neill N, Khan M, Kerwan A, Al-Jabir A, et al. World Health Organization declares global emergency: A review of the 2019 novel coronavirus (COVID19). International Journal of Surgery. 2020; 76: 71–76

4. Worldometer on coronavirus disease 2019. https://www.worldometers.info/coronavirus/

5. World Health Organization. Coronavirus disease 2019 (COVID-19): situation report, 72. 2020.

6. Ozili PK. COVID-19 in Africa: socioeconomic impact, policy response, and opportunities. Policy Response and Opportunities (April 13, 2020). 2020.

7. Ataguba JE. COVID-19 pandemic, a war to be won: understanding its economic implications for Africa. Appl Health Econ Health Policy. 2020.

8. Mudenda S. Letter to Editor: Coronavirus Disease (COVID-19): A Global Health Problem. Int J Pharm Pharmacol 2020; 4(1):141

9. Zambia National Public Health Institute (ZNPHI). http://znphi.co.zm/news/wpcontent/uploads/2020/03/Ministerial-statement_COVID-19-SI_14Mar2020_min.pdf

10. International Pharmaceutical Federation (FIP). SRAS-CoV-2 outbreak preparedness. https://www.fip.org/coronavirus. Accessed 19 Apr 2020.

11. Kalungia AC, Burger J, Goodman B, Costa JdO, Simuwelu C. Non-prescription sale and dispensing of antibiotics in community pharmacies in Zambia. Expert review of anti-infective therapy. 2016; 14(12):1215–1223

12. Phiri MN, Banda M, Mudenda S, et al. Coronavirus Disease 2019 (COVID-19): The Role of Pharmacists in the Fight against COVID-19 Pandemic. Int J Pharm Pharmacol. 2020; 4(1):143

13. Cadogan CA, Hughes CM. On the frontline against COVID-19: Community pharmacists’ contribution during a public health crisis. Research in Social and Administrative Pharmacy. 2020.

14. Wang CJ, Ng CY, Brook RH. Response to COVID-19 in Taiwan: big data analytics, new technology, and proactive testing. JAMA. 2020.

15. Carico R, Sheppard J, Thomas CB. Community pharmacists and communication in the time of COVID-19: Applying the health belief model. Research in Social and Administrative Pharmacy. 2020.

16. Putt W. Health Care Policy: Walgreens, CVS Health Step Up Buyout Offers During Pandemic. 2020. https://www.truthrx.org/puttnewswire/health-care-policy-walgreens-cvs-health-step-up-buyout-offers-during-pandemic

17. Amariles P, Ledezma-Morales M, Salazar-Ospina A, Hincapié-García JA. How to link patients with suspicious COVID-19 to the health system from the community pharmacies? A route proposal. Research in Social and Administrative Pharmacy. 2020; S1551-7411(20)30248-5

18. Adunlin G, Murphy PZ, Manis M. COVID-19: How Can Rural Community Pharmacies Respond to the Outbreak? The Journal of Rural Health. 2020

19. Zambia Medicines Regulatory Authority (ZAMRA). http://www.zamra.co.zm/wp-content/uploads/2019/07/REGISTER-OF-CERTIFICATES-OF-REGISTRATION.pdf

20. Hui DS, Azhar E, Madani TA, Ntoumi F, Kock R, Dar O, Ippolito G, Mchugh TD, Memish ZA, Drosten C. The continuing 2019-nCoV epidemic threat of novel coronaviruses to global health-the latest 2019 novel coronavirus outbreak in Wuhan, China. Int. J. Infect. Dis. 2020; 91: 264–266

21. World Health Organization. Infection prevention and control of epidemic- and pandemic-prone acute respiratory infections in health care [EB/OL]. [2020-02-20]. https://apps.who.int/iris/bitstream/handle/10665/112656/9789241507134_eng.pdf;jsessionid=2951A95A10A851883E036F6869AE985B?sequence=1.

22. Ung COL. Community pharmacist in public health emergencies: quick to action against the coronavirus 2019-nCoV outbreak. Res Social Adm Pharm. 2020; 16(4):583–586

23. Special Committee for Drug Borne Diseases of Chinese Pharmacology Society. Self-management strategies for home-living elderly patients on multiple drugs for treating chronic disease [EB/OL]. [2020-02-20]. https://mp.weixin.qq.com/s/YpCTEWvPcHdPtVnTK41mjw.

24. Liu S, He G, Du J, Wang D, Shi C, Huang Q, et al. Pharmaceutical emergency guarantee difficulties and countermeasures for the prevention and control of an outbreak of novel coronavirus pneumonia (NCP). Chin J Hosp Pharm. 2020; 40(3):243–249

25. Zheng S-Q, Yang L, Zhou P-X., Li H-B., Liu F, Zhao R-S. Recommendations and guidance for providing pharmaceutical care services during COVID-19 pandemic: A China perspective. Research in Social and Administrative Pharmacy. 2020. https://doi.org/10.1016/j.sapharm.2020.03.012

26. Wei M, Chen Q, Yu DM, et al. The development of pharmaceutical care in domestic and foreign communities and its enlightenment on establishing pharmacy major in China [J]. Chin Pharmaceut J. 2010; 12: 98–100

27. Centers for Disease Control and Prevention. Preventing COVID-19 spread in communities [EB/OL]. [2020-03-04]. https://www.cdc.gov/coronavirus/2019-ncov/community/index.html.

28. World Health Organization. Coronavirus disease (COVID-19) advice for the public: when and how to use masks [EB/OL]. [2020-03-04]. https://www.who.int/emergencies/diseases/novel-coronavirus-2019/advice-for-public/when-and-how-touse-masks.

29. Chinese Academy of Medical Science Peking Union Medical College. Peking Union Medical College Novel Coronavirus Infection Prevention Q&A for the Public [EB/OL]. Beijing: Publishing House of Peking Union Medical College; 2020 [2020-02-20]. https://ncovh5.yibaomd.com/ncov/v2?from=singlemessage&isappinstalled=0.

30. Basheti IA, Nassar R, Barakat M, Alqudah R, Abufarha R, Mukattash TL, Saini B. Pharmacists’ readiness to deal with the coronavirus pandemic: Assessing awareness and perception of roles. Research in Social and Administrative Pharmacy. 2020.

